# Evaluating MRI harmonisation methods with multicentre data from the Psy-ShareD consortium: A comparative analysis

**DOI:** 10.64898/2026.01.12.26343904

**Authors:** Mariana Zurita, Rubaida Easmin, Stephen Lawrie, Heather Whalley, Aleks Stolicyn, Jane Garrison, Graham K. Murray, Shun-Chin Jim Wu, Tsutomu Takahashi, Giuseppe Pontillo, Felice Iasevoli, Urvakhsh M. Mehta, Rachel Upthegrove, Sophia Frangou, Simon Evans, Veena Kumari, Jack Rogers, Matthew J Kempton, Paul Allen, Psy-ShareD Partnership

## Abstract

Combining multi-site MRI datasets increases statistical power and model generalisability but may be hindered by variability between sites. Harmonisation methods aim to remove potentially confounding variance while preserving biologically meaningful signals. However, this can be challenging, as each T1-weighted image reflects both scanner properties (e.g., field strength, sequence parameters) and individual biological characteristics (e.g., age, sex, ethno-cultural background, and pathology). Two image-based (HACA3, IGUANe) and two feature-based (neuroHarmonize, neuroCombat) harmonisation methods were assessed using T1-weighted brain imaging data from the Psy-ShareD database; 564 participants (295 schizophrenia, 269 controls) from seven studies acquired across 5 sites from the Psy-ShareD database. We trained several models to classify sites, schizophrenia diagnosis, age, and symptom levels. Site-classification accuracy was high for unharmonised data (90.1%) and for HACA3 (92.2%), slightly reduced with IGUANe (86.6%), and near chance for feature-based methods (4.2% neuroHarmonize; 1.8% neuroCombat), indicating effective bias removal. We fitted several models predicting biological signals including diagnosis, age, and symptom levels across different harmonisation methods. In most cases, classification with harmonised data performed at least as well as with unharmonised data. Generally, feature-based methods best remove site-related variance, but image-based approaches remain a promising avenue for preserving individual biological differences. This work provides practical guidance for selecting harmonisation strategies in multi-site psychiatric neuroimaging, depending on whether the priority is bias reduction or preservation of subject-level variability.

## 1. Introduction

Magnetic Resonance Imaging (MRI) is a crucial tool for studying the brain’s structure, with T1-weighted imaging being one of the most widely used modalities due to its ability to provide high resolution spatial information on gray and white matter structures. However, the reproducibility and generalisability of MRI-derived measurements across different scanners, site and acquisition protocols remain a significant challenge in neuroimaging research (Pomponio et al., 2020; Roca et al., 2025). Variations in scanner hardware, acquisition parameters, and preprocessing pipelines introduce differences in the data (Biberacher et al., 2016, Hedges et al., 2022), generating site-effects, which are known to affect the ability to derive useful conclusions from the data (Johnson et al., 2007). These differences lead to biased and inconsistent measurements of true biological effects of interest (Biberacher et al., 2016; Wen et al., 2016; Zuo et al., 2023), which can compromise the validity of multi-site studies or meta-analyses.

Another challenge in MRI studies is the limited availability of large and ethnoculturally diverse samples due to the high costs and logistical challenges of scanning. It is also more difficult to recruit individuals with psychiatric disorders, such as schizophrenia, across different sites, which further complicates the analyses of multi-site datasets (Evans & Allen, 2024). To address these issues, the Psychosis MRI Shared Data Resource (Psy-ShareD; https://psyshared.com/) was created to pool neuroimaging data from sites worldwide, increasing the sample size and enhancing the potential generalisability of analyses and findings across different populations and scanner protocols (Allen et al., 2024). However, site-to-site differences in scanner parameters and acquisition protocols still remain an issue.

Harmonisation methods have been developed to address these challenges, and reduce the impact of site-related variability, enabling more accurate cross-site comparisons and enhancing the statistical power of studies involving pooled data (Fortin et al., 2018; Johnson et al., 2007; Roca et al., 2023; Wen et al., 2023; Zuo et al., 2023). Applied to MRI data, these methods can be broadly classified into feature-based, or implicit methods, and image-based, or explicit methods (Wen et al., 2023; Zuo et al., 2023). Traditional feature-based methods (Wen et al., 2023) such as ComBat (Fortin et al., 2018) usually involve applying statistical adjustments, to remove site effects from features extracted from the image, such as cortical thickness (Roca et al., 2023). More recent image-based methods, on the other hand, are a specific type of image-to-image translation method (Zuo et al., 2023; Roca et al., 2025). These methods use models that take the original unharmonised image as input and output a harmonised image, while preserving the original image’s anatomical features (Zuo et al., 2023; Wen et al., 2023).

Several types of models are used for image-based harmonisation, with most relying on deep-learning methods such as generative adversarial networks (GAN; Roca et al., 2023), autoencoders (Zuo et al., 2023), and convolutional neural networks (Wen et al., 2023). The choice of harmonisation method can significantly influence the outcomes of neuroimaging studies, making it crucial to understand the strengths and limitations of each approach (Wen et al., 2023; Zuo et al., 2023).

Here we provide a comparative analysis of four different harmonisation methods (IGUANe, HACA3, neuroHarmonize and neuroCombat, defined below) applied to T1-weighted brain MRI data from PsyShareD database, in individuals with schizophrenia and healthy controls. We examined the performance of these methods in terms of their ability to mitigate scanner-related variability, preserve biological signals, and maintain the interpretability of the results. By highlighting the advantages and potential limitations of each method, we aim to provide a valuable guide for researchers seeking to optimise the accuracy and reliability of multi-site neuroimaging studies.

## 2. Methods

This section details the data sources, harmonisation methods, image processing pipeline, and statistical analyses used in this study. We first describe the two datasets used: the primary dataset from Psy-ShareD and a secondary dataset from the Brain Segmentation Testing Protocol. Next, we outline the four harmonisation methods applied to all T1-weighted images, including two image-based methods and two feature-based methods. We then describe how FreeSurfer was used to extract structural features from both the original and harmonised images. Following this, we present the analytical approaches used to evaluate the effects of harmonisation. These include a site-classification task to quantify the removal of site-related variance, and a series of models assessing whether biologically and clinically meaningful signals—such as diagnosis, age, and symptom severity—were preserved. Finally, we describe the analysis conducted on the second dataset to assess the consistency of volume estimates across scans with different acquisition parameters.

### 2.1. Data

#### 2.1.1. Psy-ShareD data

Data for the first part of this study were obtained from Psy-ShareD (https://psyshared.com/), an open-access global repository of multi-site T1-weighted MRI data collected from individuals with schizophrenia spectrum disorders, individuals at clinical high risk for psychosis, and healthy controls (Allen et al., 2025). For the present analysis, we included data from seven studies that had balanced numbers of individuals with a schizophrenia diagnosis and healthy controls (Cooke et al., 2008; Barkataki et al., 2006; Garrison et al., 2017; Chuang et al., 2014; Cooper et al., 2023; Tranfa et al., 2023; Sasabayashi et al., 2021). These studies were conducted across five sites (university, ID: King’s College London, 01; University of Cambridge, 04; University of Edinburgh, 07; University of Naples Federico II, 19; University of Toyama, 21) in three countries (United Kingdom, Italy and Japan), comprising 295 individuals diagnosed with schizophrenia and 269 healthy controls (Table 1). Within each study, the same imaging protocol and scanner were used for all participants. Age and symptom severity scores differed significantly across sites (both ANOVA p < 0.001), underscoring the importance of harmonisation methods that preserve clinically relevant variance (Figure 1A).

**Table 1:** Description of data used [HC: healthy controls; SCZ: people with schizophrenia].

| Dataset | Participants<br>HC / SCZ | Age ± SD<br>HC / SCZ | Sex<br>HC / SCZ | Scanner | PSYD<br>Site ID | Location |
| --- | --- | --- | --- | --- | --- | --- |
| PSYD_0106 | 31 / 66 | 33.5 ± 11.4 /<br>38.0 ± 9.6 | M: 22, F: 9 /<br>M: 49, F: 17 | 1.5T GE | 01 | London,<br>UK |
| PSYD_0118 | 15 / 28 | 32.1 ± 7.2 /<br>34.5 ± 6.2 | M: 15, F: 0 /<br>M: 28, F: 0 | 1.5T GE | 01 | London,<br>UK |
| PSYD_0401 | 20 / 20 | 33.4 ± 7.9 /<br>36.3 ± 7.2 | M: 18, F: 2 /<br>M: 18, F: 2 | 3T<br>Siemens | 04 | Cambridge,<br>UK |
| PSYD_0403 | 21 / 21 | 34.3 ± 9.9 /<br>32.2 ± 7.3 | M: 17, F: 4 /<br>M: 18, F: 3 | 3T<br>Siemens | 04 | Cambridge,<br>UK |
| PSYD_0701 | 40 / 34 | 37.7 ± 14.3 /<br>36.6 ± 10.1 | M: 21, F: 19 /<br>M: 24, F: 10 | 3T<br>Siemens | 07 | Edinburgh,<br>UK |
| PSYD_1901 | 55 / 49 | 42.4 ± 15.6 /<br>37.5 ± 9.6 | M: 29, F: 26 /<br>M: 34, F: 15 | 3T<br>Siemens | 19 | Naples,<br>Italy |
| PSYD_2101 | 87 / 77 | 26.3 ± 3.9 /<br>28.8 ± 9.4 | M: 46, F: 41 /<br>M: 39, F: 38 | 3T<br>Siemens | 21 | Toyama,<br>Japan |

**Figure 1:**
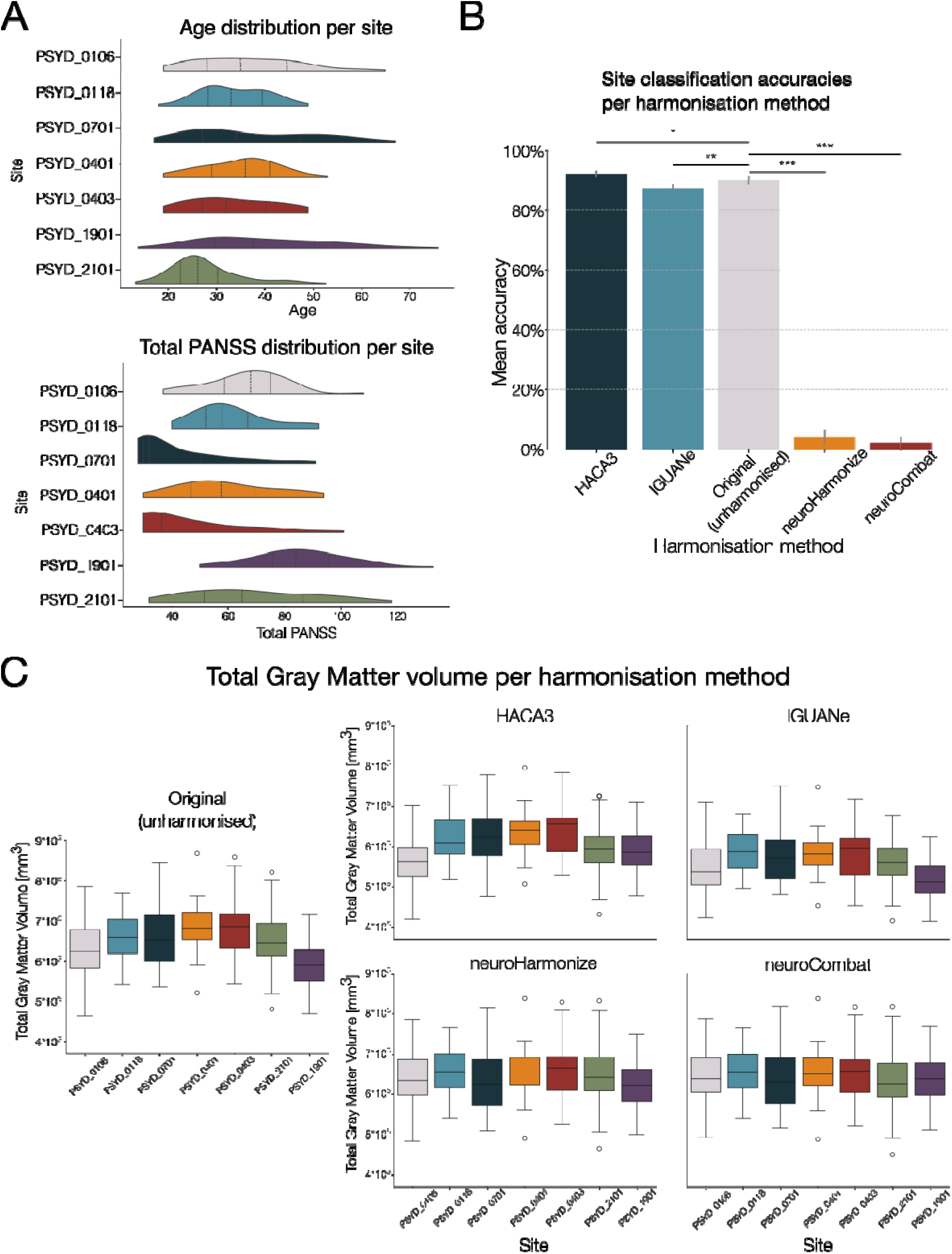
Dataset characteristics and harmonisation effects. (A) Age and total PANSS scores distributions per site. (B) Site-classification accuracy according to harmonisation method. [n.s.: non-significant; *: p<0.05; **: p<0.01; ***p<0.001] (C) Total Gray Matter volume values per site, per harmonisation method.

#### 2.1.2. Brain Segmentation Testing Protocol data

The gold standard for evaluating harmonisation performance is the inclusion of travelling subjects. Since this was not available in the PsyShareD dataset (as would likely be the case for studies with retrospectively acquired data), for the final analysis of the study we used a separate dataset from the Brain Segmentation Testing Protocol, a publicly available collection of MRI images designed for evaluating segmentation algorithms (Kempton et al., 2011; https://sites.google.com/site/brainseg). This dataset includes 72 images from 9 adults (age = 28 ± 8.5 years, 6 females), each of whom was scanned using two MRI scanners (1.5T and 3T General Electric Signa HDx) with four different pulse sequences per scanner (8 images per subject in total; mean interval between the scans with 1.5T and 3T scanners = 6.7 ± 4.2 days). This setup provided a unique opportunity to evaluate which harmonisation method produced the most consistent volume estimates across different image acquisitions, as any within-subject variance would most likely be due to scanner- or sequence-related differences rather than due to biological variation.

### 2.2. Image-based harmonisation

We evaluated two image-based harmonisation methods: Harmonization with Attention-based Contrast, Anatomy, and Artifact Awareness (HACA3; Zuo et al., 2023) and Image Generation with Unified Adversarial Networks (IGUANe; Roca et al., 2025). We used the preprocessing pipelines recommended by the developers of each method respectively, as these workflows are integral to how each method is intended to operate and are therefore the most representative of how researchers are likely to apply these tools in practice.

#### 2.2.1. HACA3 harmonisation

HACA3 is an unsupervised deep-learning image-to-image harmonisation method that follows an encoder–attention–decoder architecture comprising three components: (1) encoding, (2) anatomy fusion with attention, and (3) decoding (Zuo et al., 2023; https://github.com/lianruizuo/haca3). The method is autoencoder-based and aims to harmonise images by learning disentangled representations of anatomical structure, image contrast, and artefacts (Wen et al., 2023).

The encoding step includes three separate encoders (anatomy, contrast, and artefact) which allow the model to isolate these components in both input and target images. The harmonised image is generated using the anatomical information from the input image, the contrast properties from the target image, and artefact information from both images (Zuo et al., 2023). Importantly, this architecture supports within-subject harmonisation using images acquired with different sequences, enabling the anatomy encoder to integrate complementary information from multiple contrasts. The fused anatomical information is processed using attention mechanisms before being passed to the decoder, which synthesises a harmonised image that preserves subject-specific anatomy while adopting the contrast profile of the target (Zuo et al., 2023).

We followed the preprocessing steps recommended by the developers (https://github.com/lianruizuo/haca3). First, all images underwent N4 bias field correction using SimpleITK (Beare, Lowekamp, & Yaniv, 2018). For non-isotropic images, resolution was corrected using SMORE (Zhao et al., 2020). All images were then registered to MNI space using ANTsPy’s symmetric normalisation (affine and deformable transformation, with mutual information as optimisation metric; Avants et al., 2011), and the inverse transformation matrix was saved. Harmonisation was performed in MNI space using HACA3, with the pre-trained model provided in the package, and the resulting harmonised images were then transformed back into subject space using the saved inverse transformation.

#### 2.2.2. IGUANe harmonisation

IGUANe is an unsupervised, 3D image-to-image harmonisation method based on the CycleGAN framework (Roca et al., 2025; https://github.com/RocaVincent/iguane_harmonization). Its fully 3D architecture enables it to process entire brain volumes rather than image slices, allowing for more anatomically coherent translations. IGUANe adopts a many-to-one harmonisation strategy, in which forward and backward generators translate images from multiple source sites to a designated reference site and vice versa. Adversarial discriminators are used to distinguish between real and synthesised images, guiding the generators to produce realistic harmonised outputs.

First, all images were reoriented to FSL’s standard orientation to ensure consistency among sites’ Nifti headers. Then, we followed the preprocessing pipeline recommended by the developers. Skull-stripping was performed using HD-BET (Isensee et al., 2019), followed by N4 bias field correction using N4ITK (Tustison et al., 2010). Images were then registered to MNI space using FSL-FLIRT (Jenkinson et al., 2002), cropped to 160 × 192 × 160 voxels, and normalised by dividing by the median intensity within the brain mask.

The preprocessed images were then harmonised using IGUANe, employing pretrained model parameters provided by the authors of the method. After harmonisation, output images were post-processed by padding them to their original dimensions and applying the inverse transform to return them from MNI space to subject space.

### 2.3. Feature-based harmonisation

For feature-level harmonisation, we selected two commonly used methods: neuroCombat (Fortin et al., 2018) and neuroHarmonize (Pomponio et al., 2020). Both methods operate on extracted image features (i.e., cortical thickness values) and aim to remove site-related variability while preserving biologically meaningful variation.

#### 2.3.1. neuroCombat

neuroCombat is a harmonisation tool that adjusts for scanner- and site-related variability in neuroimaging features (Fortin et al., 2018; https://github.com/Jfortin1/neuroCombat). It is an extension of the ComBat method originally developed for genomics and later adapted to neuroimaging data (e.g., diffusion tensor imaging). The method applies a linear model to remove site effects, while preserving variance associated with biological covariates. It accounts for site-specific scaling and location parameters, and uses empirical Bayes estimation to stabilise parameter estimates, particularly in datasets with smaller sample sizes per site.

In all neuroCombat analyses, we included sex and age as biological covariates, except in the age classification task, where age was the target variable (details provided in the relevant Results section). This was done to preserve the biologically relevant variance in the data and only correct for site effects. Although ComBat-like harmonisation frameworks allow inclusion of multiple covariates, we did not include PANSS or diagnosis as covariates because doing so would remove variance directly related to psychiatric pathology, which is one of the primary biological signals we aimed to evaluate downstream.

#### 2.3.2. neuroHarmonize

neuroHarmonize extends the ComBat framework by incorporating generalised additive models (GAMs) to model non-linear age effects more flexibly (Pomponio et al., 2020; https://github.com/rpomponio/neuroHarmonize). Specifically, it adds a penalised non-linear term to estimate the relationship between age and ROI volumes, allowing the harmonisation process to adapt to non-linear developmental or degenerative patterns (Pomponio et al., 2020). This approach is particularly advantageous in lifespan or heterogeneous datasets where age-related trajectories are not expected to be linear.

As with neuroCombat, we included sex and age as covariates in all neuroHarmonize analyses, except in the age classification task, where harmonisation was performed without age as a covariate. In that case, the two harmonisation methods were effectively equivalent, as both defaulted to ComBat without age adjustment.

### 2.4. Image processing

Harmonised and unharmonised images were processed using FreeSurfer version 7.3.2 (surfer.nmr.mgh.harvard.edu) using the defaul*t recon-all* command to extract cortical and surface measures (number of vertices, surface area, gray matter volume, cortical thickness, and curvature), particularly from recon-all’s outputs brainvol.stats and aparc.DKatlas files. These are the values used for all the following analyses.

### 2.5. Harmonisation methods comparison

#### 2.5.1. Site-classification

One of the primary goals of harmonisation was to reduce site-related bias. To evaluate this, we trained a multi-class Support Vector Machine (SVM) classifier using the scikit-learn package in Python (Pedregosa et al., 2011). Each model was trained to predict the acquisition site based on FreeSurfer-derived features. Separate SVM models were trained on the original (unharmonised) data and on the data harmonised using each of the four methods described above. We used a leave-one-site-out cross-validation to assess model performance in each case. With this approach, a reduction in classification accuracy after harmonisation indicates successful removal of site effects.

#### 2.5.2. Models assessing preservation of biological signal

To assess whether harmonisation preserved meaningful biological variation, we evaluated model performance across three prediction targets: diagnosis, age, and symptom assessment scores using the Positive and Negative Syndrome Scale (PANSS; Kay, Fiszbein, & Opler, 1987). If harmonisation successfully preserves biological and clinical signals, model performance should be at least comparable to that of the original data across all prediction targets. All the models were built using the scikit-learn package in Python (Pedregosa et al., 2011) and tested for different numbers of features using leave-one-site-out cross-validation.

##### Diagnosis classification

To test whether disease-related signals were preserved, we trained binary linear SVM classifiers to distinguish individuals with schizophrenia from healthy controls. SVM classifiers were trained using nested cross-validation, in which the outer loop used leave-one-study-out cross-validation to simulate generalisation across sites. The inner loop applied recursive feature elimination (RFE) to select the optimal features to improve the model accuracy.

##### Age classification

One of the most noticeable effects of aging on the brain structure is the reduction in its volume, primarily due to the loss of gray matter and the enlargement of ventricles (Lee & Kim, 2022). To further assess the preservation of biological signals, we examined whether harmonised data retained information about age at a broad level. We performed a mean-split age classification in healthy controls, dividing participants into “younger” and “older” groups based on the overall sample mean (34 years). To sharpen the contrast between groups, 27 participants aged between 33 and 35 were excluded.

We trained logistic regression models with L1-based feature selection to classify age group membership. As with diagnosis classification, we used study-level cross-validation, where each fold left one study out for testing. We compared performance across harmonisation methods using the same linear mixed-effects framework described below.

##### PANSS prediction

PANSS scores have been associated with cortical thinning, with positive and total symptom scores linked to regional reductions in cortical thickness, and negative symptoms associated with widespread thinner cortex in both hemispheres (Van Erp et al., 2018). To evaluate whether harmonisation methods preserved clinically relevant information in patient data, we used linear regression models to predict PANSS subscale scores. Separate models were trained to predict scores on the general, positive, and negative PANSS symptom subscales.

The model pipeline included a nested cross-validation, with the outer loop consisting of a leave-one-study-out cross-validation, and the inner loop used for feature selection using scikit-learn package’s SelectKBest function with the f_regression criteria (Pedregosa et al., 2011). The linear regression model performance was evaluated using the mean absolute error (MAE) between predicted and actual PANSS subscale scores was used to assess performance.

We used linear mixed-effects models to statistically compare harmonisation methods (please see the section below for further details). The dependent variable was the MAE, with harmonisation method as a fixed effect and cross-validation fold as a random intercept.

##### Model results comparison

To compare performance of the models assessing preservation of biological signals across harmonisation methods, we calculated each model’s accuracy, precision, recall and mean absolute error (MAE). We then fit linear mixed-effects models using the ‘statsmodels’ package (Seabold and Parktold, 2010). The dependent variable was the classification metric of interest, with harmonisation method as a fixed effect and number of features as a covariate. Cross-validation fold identity was included as a random intercept to account for repeated measures. By re-levelling the harmonisation factor to set ‘original’ as the reference category, we could directly test whether each harmonisation method significantly differed from the unharmonised baseline using Wald z-tests.

#### 2.5.3. Same-subject, different sequence analysis

To evaluate whether harmonisation improved consistency across scans acquired with different imaging parameters, we applied each harmonisation method to the Brain Segmentation Testing Protocol dataset (Kempton et al., 2011; https://sites.google.com/site/brainseg), described above. For each participant, we extracted total gray matter volume from all eight scans and computed the standard deviation across these values, reflecting within-subject variability, for each harmonisation method. Lower variability indicates greater consistency across sequences. We used Wilcoxon signed-rank tests to compare within-subject variability in the harmonised data against that in the original (unharmonised) images.

## 3. Results

### 3.1. Site-classification

A core goal of harmonisation is to reduce or eliminate site-related bias. To quantify this, we trained a multi-class support vector machine (SVM) model to classify the site of origin based on FreeSurfer-derived features. As would be expected, using the original (unharmonised) data, the model achieved 90.1% ± 1.3% accuracy (Figure 1B), indicating strong site-specific signatures.

On the one hand, the image-level harmonisation methods showed mixed performance. The HACA3 model increased site classification accuracy to 92.2% ± 1.3%. A paired t-test comparing the accuracies for each site confirmed that this was significantly higher than in the unharmonised data (t = –2.3, p = 0.048), suggesting that HACA3 may have inadvertently amplified site-specific differences. In contrast, IGUANe reduced site classification accuracy to 86.6% ± 1.5%, a significant improvement relative to the original data (t = 3.4, p = 0.009), indicating a reduction in site-related variance.

On the other hand, feature-level harmonisation methods were highly effective at eliminating site effects. Relative to unharmonised data, neuroCombat reduced site classification accuracy to 1.8% ± 1.7% (t = 82.1, p < 0.001), and neuroHarmonize achieved 4.2% ± 2.9% (t = 54.3, p < 0.001). These results indicate that feature-based methods nearly abolished site identifiability, which is encouraging in terms of reducing site related bias. However, it raises concern that biologically meaningful variance might also be removed in the process, despite inclusion of biological covariates during correction. To illustrate this, Figure 1C shows boxplots of total gray matter volume per site for each harmonisation method. While image-level methods preserved some inter-site variability—likely reflecting real demographic or clinical differences across samples (see Figure 1A)—feature-level methods dramatically compressed this variance, potentially at the cost of losing meaningful biological signal.

We therefore tested whether key biological characteristics, such as diagnosis, age and symptom severity, could still be identified in the harmonised data, and how prediction performance compared to the original dataset.

### 3.2. Models assessing preservation of biological signal

To evaluate whether the harmonisation preserved biologically relevant signals, we conducted three separate analyses: (1) diagnosis classification; (2) age classification; (3) PANSS subscale score prediction.

#### Diagnosis classification

The aim of the diagnosis classification analysis was to determine whether harmonisation supported the detection of biologically meaningful features that distinguish individuals with schizophrenia from healthy controls across the 7 Psy-ShareD datasets. Given that support vector machine (SVM) performance is sensitive to the number of input features, we conducted the analysis across several feature set sizes (Wang et al., 2024). Classification with each harmonisation method performed at least as well as with the original (unharmonised) data, indicating no detrimental effect of harmonisation on classification performance (Figure 2; left column).

**Figure 2:**
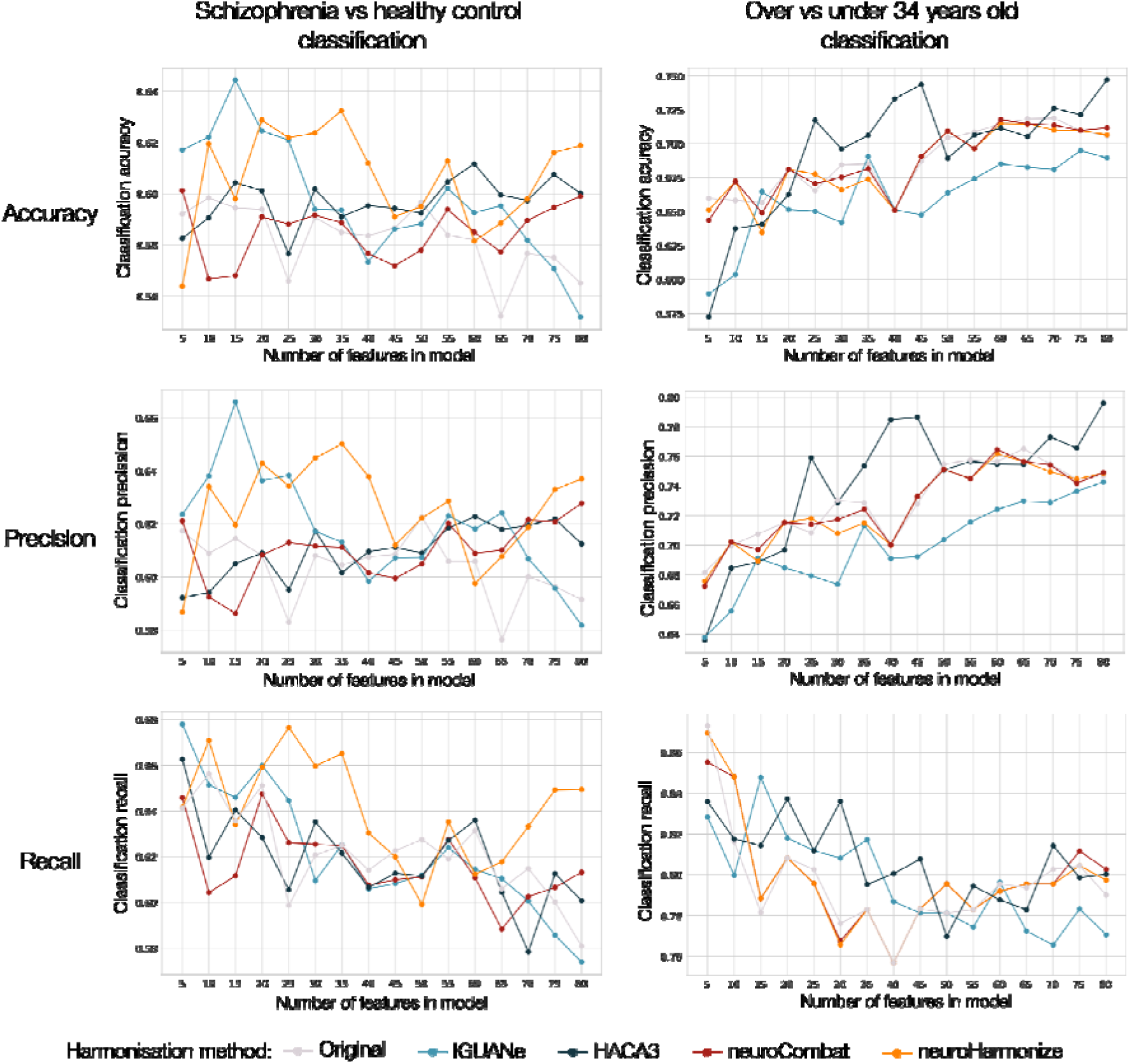
Classification model results. *Left column:* Mean results from SVM models that classify between individuals with schizophrenia and healthy controls for different numbers of features permitted in the model. *Right column:* Mean results from logistic regressions models’ classification of older vs younger than 34 year-old healthy controls, for different maximum numbers of features in the model. The results are presented in three columns, for accuracy, precision and recall plots, respectively.

To formally assess the impact of harmonisation, we fitted linear mixed-effects models predicting classification accuracy, precision, and recall, with harmonisation method as a fixed effect and number of features as a covariate (Supplemental Table 2). Compared to the original data, accuracy was significantly improved with HACA3 (β = 0.014, p = 0.047), IGUANe (β = 0.015, p = 0.040), and neuroHarmonize (β = 0.024, p = 0.001), while application of neuroCombat showed no significant accuracy difference (β = 0.002, p = 0.734). Overall, neuroHarmonize provided the most substantial improvement across classification metrics. In contrast, neuroCombat, despite its effectiveness in removing site-specific variance, did not enhance diagnostic classification performance.

#### Age classification

We then assessed age classification performance within the healthy control group as a benchmark for biologically grounded signal, using a binary logistic regression to distinguish participants as younger or older than the mean sample age (34 years). Accuracy, precision, and recall values for classifications with each harmonisation method across different feature set sizes are shown in Figure 2 (right column).

To quantify the effects of harmonisation on classification performance, we fit linear mixed-effects models predicting each performance metric (accuracy, precision, and recall) as a function of harmonisation method, with the number of features included as a covariate and the unharmonised dataset serving as baseline (Supplemental Table 3). The model revealed that classification with HACA3, neuroHarmonize and neuroCombat did not differ significantly from classification with the original data in terms of accuracy (HACA3: β = 0.007, p = 0.450; neuroHarmonize: β = –0.003, p = 0.734; neuroCombat: β = –0.001, p = 0.884), meaning that these methods preserved the biologically-relevant information for predicting age. IGUANe, by contrast, significantly decreased classification accuracy (β = –0.028, p = 0.002). These findings suggest that, in contrast to diagnosis classification, harmonisation may not improve the detection of biologically meaningful age-related differences in healthy individuals, and in the case of IGUANe, it may hinder the predictive performance.

#### PANSS score prediction

To evaluate whether harmonisation preserved clinically relevant information, we focused on the individuals with schizophrenia and assessed the ability of linear models to predict symptom severity, specifically the General, Negative, and Positive PANSS subscale scores. For each score, we trained regression models and computed the mean absolute error (MAE) between the predicted and actual values, as shown in Figure 3. We then used linear mixed-effects models to predict MAE based on the harmonisation method and number of features, with the original unharmonised data serving as the baseline (results in Supplemental Table 4). For PANSS General subscale scores, HACA3 significantly reduced the prediction error (β = –0.179, p = 0.015), while IGUANe (β = –0.096, p = 0.193), neuroHarmonize (β = 0.030, p = 0.682), and neuroCombat (β = –0.049, p = 0.511) did not differ significantly from the original data.

**Figure 3:**
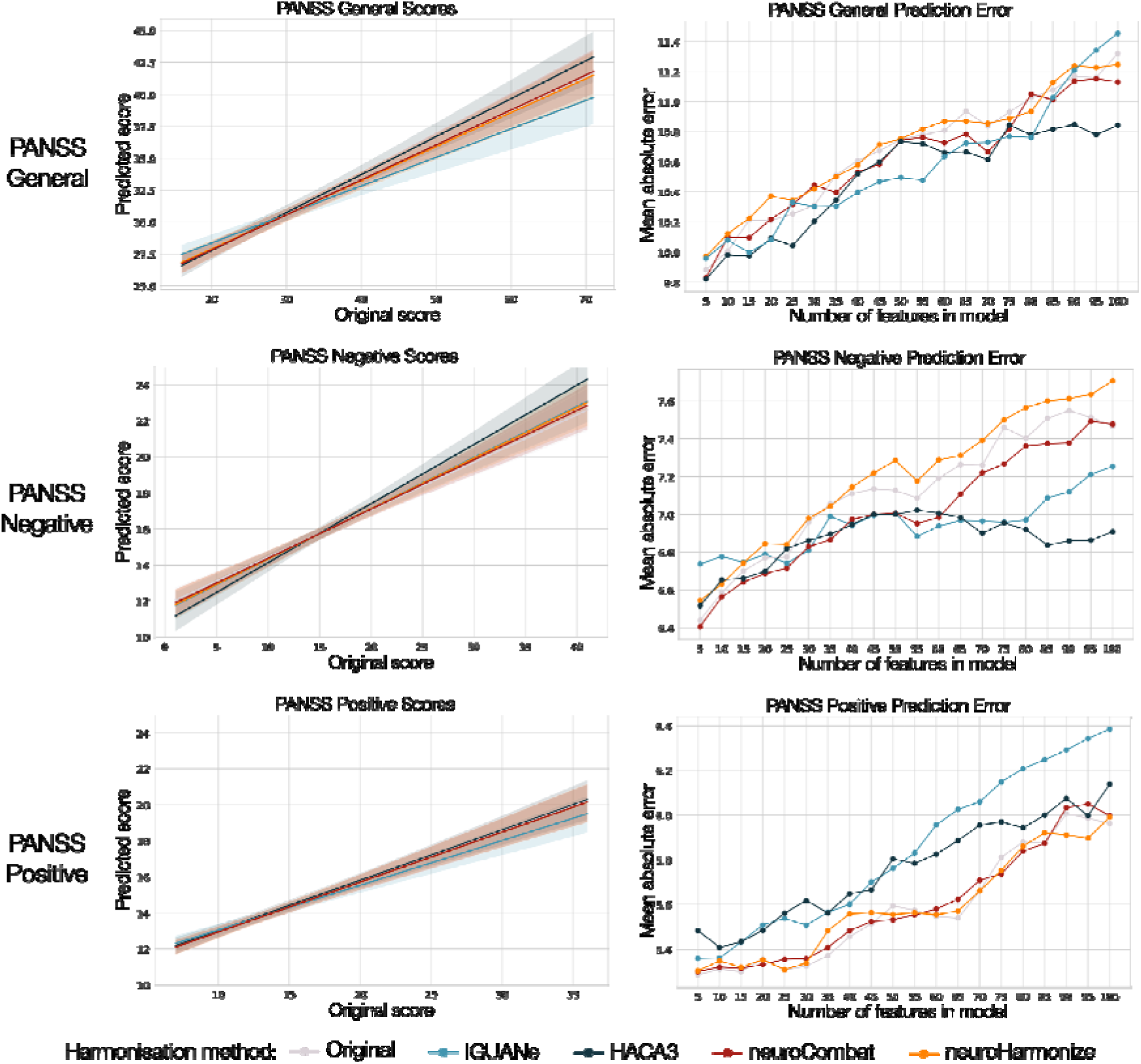
PANSS subscale score prediction models. *Left column:* Linear regressions showing the relationship between the original scores and the PANSS subscale scores predicted by the linear regression models trained with harmonised FreeSurfer data. *Right column:* Mean absolute error of the predicted PANSS subscale score compared to the original score, for different numbers of features in the regression model. Models trained with feature-level data generally performed similarly to those trained with unharmonised data.

**Figure 4:**
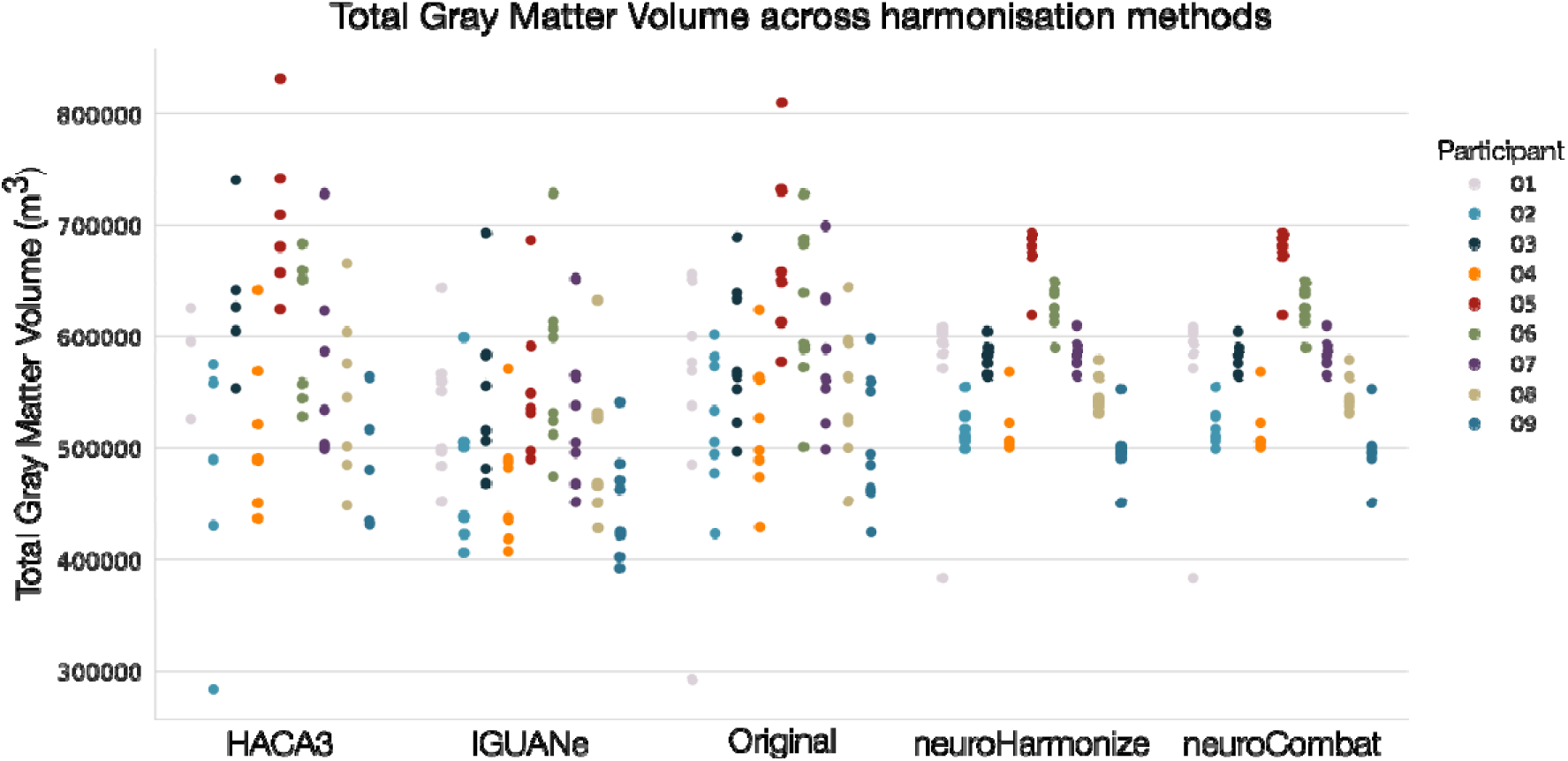
Harmonised values for GM volume. Gray Matter (GM) volumes for nine subjects scanned across two scanners with eight different sequences for different harmonisation methods. Image-level harmonisation (HACA3 and IGUANe) produced a high variance in GM volumes, while feature-level harmonisation (neuroHarmonize and neuroCombat) reduced it.

For PANSS Negative subscale scores, both HACA3 (β = –0.250, p < 0.001) and IGUANe (β = –0.174, p < 0.001) outperformed prediction with unharmonised data, whereas neuroCombat also improved performance but more modestly (β = –0.103, p = 0.009). In contrast, neuroHarmonize slightly worsened prediction accuracy (β = 0.085, p = 0.031). For PANSS Positive subscale scores, none of the methods improved performance, and both HACA3 (β = 0.179, p < 0.001) and IGUANe (β = 0.258, p < 0.001) significantly increased prediction error, while neuroHarmonize (β = 0.008, p = 0.826) and neuroCombat (β = 0.013, p = 0.709) performed similarly to the original data. These findings suggest that while certain harmonisation methods may enhance prediction of specific symptom dimensions, particularly negative symptoms, others may compromise the sensitivity to variation in positive symptoms.

### 3.3. Same-subject, different sequence analysis

Finally, to provide an independent assessment of consistency across harmonisation methods, we conducted a separate analysis using data from nine individuals scanned under four different sequences on two different scanners, resulting in eight scans per subject. As we would expect biological variance to be small within each subject, this allowed us to test the effect of the harmonisation methods on scanner related variance. This dataset allowed us to examine how each harmonisation method affected within-subject variability in Total Gray Matter Volume. Using the Wilcoxon signed-rank test to compare each harmonised dataset’s variance to the original, we found that both neuroHarmonize and neuroCombat significantly reduced within-subject variance (p = 0.0039), with an average reduction of 81.8%. Note that in this analysis neuroHarmonize is equivalent to the neuroCombat processing, as we omitted the age covariate. In contrast, application of IGUANe and HACA3 did not result in a statistically significant change (IGUANe: p = 0.359; HACA3: p = 0.426). These findings suggest that feature-level harmonisation methods, particularly neuroHarmonize and neuroCombat, improve measurement consistency across repeated scans, reinforcing their utility in mitigating non-biological variability.

## 4. Discussion

In this study, we systematically compared four harmonisation methods (HACA3, IGUANe, neuroCombat, and neuroHarmonize) applied to multi-site structural MRI data from the Psy-ShareD resource and a separate same-subject test dataset from the Brain Segmentation Testing Protocol. Our goal was to evaluate their ability to reduce site-related variance while preserving biologically meaningful signals. Overall, feature-level harmonisation methods, particularly neuroHarmonize, were most effective at reducing site effects without impairing downstream analyses, except in the case of PANSS Negative subscale score prediction. Image-level methods showed more variable performance, with HACA3 and IGUANe yielding improvements for some clinical signals.

A primary objective of harmonisation is to mitigate site-related variance without compromising biological signal. Our site-classification analysis revealed clear differences across methods in their ability to remove site-specific information. Feature-level approaches substantially reduced site identifiability, with classification accuracies approaching zero. This suggests that these methods successfully were effective at attenuating scanner-related variance. However, such pronounced reductions may risk removing meaningful biological differences if site effects are partially confounded with demographic or clinical variables. For instance, both age and PANSS subscale scores differed significantly across sites (both p < 0.001), and such biologically relevant differences may partly contribute to site-specific signatures. Recent developments (e.g., Gardner et al., 2025) propose modified ComBat frameworks that explicitly preserve variance in selected clinical dimensions.

Image-level methods showed more variable performance. IGUANe moderately reduced site classification accuracy, indicating partial removal of site effects, whereas HACA3 unexpectedly increased site identifiability, suggesting a possible amplification of site-specific artefacts. To further probe the impact of harmonisation on non-biological variability, we examined within-subject consistency using repeated scans from individuals scanned under different protocols. Consistent with our multiple-site analysis, feature-level methods outperformed image-level approaches: both neuroCombat and neuroHarmonize significantly reduced within-subject variance in total gray matter volume, indicating improved measurement correspondence across sequences. IGUANe showed a modest, non-significant reduction, while HACA3 slightly increased within-subject variance. These findings reinforce the robustness of feature-level harmonisation, particularly neuroHarmonize, in removing spurious variance both across and within sites, a critical requirement for multi-site neuroimaging research.

In terms of clinical utility, harmonisation methods varied in their ability to support the detection and prediction of schizophrenia-related clinical features. For diagnosis classification, neuroHarmonize provided the most consistent improvements across accuracy, precision, and recall. IGUANe and HACA3 also yielded selective gains in classification accuracy and precision, while neuroCombat resulted in no significant benefits despite effectively removing site-related variance. When predicting PANSS symptom severity, image-level methods significantly improved prediction of negative symptoms, with HACA3 also enhancing prediction of general symptoms. However, both increased error in positive symptom prediction, suggesting a trade-off in their ability to preserve different clinical signals. In contrast, neuroHarmonize and neuroCombat maintained stable performance across PANSS domains but offered no improvement as compared to the original data.

These results highlight the challenge of preserving diverse clinical signals through harmonisation and point to neuroHarmonize as the most consistent method for data harmonisation for diagnostic classification, while image-level approaches may better capture certain symptom dimensions. To evaluate the preservation of other biologically meaningful signals, we examined age classification within healthy controls. Most harmonisation methods (HACA3, neuroCombat, and neuroHarmonize) resulted in comparable performance to the original data, suggesting they preserved age-related variance. In contrast, IGUANe significantly reduced both accuracy and precision, indicating a loss of biologically relevant information. These results again underscore that not all harmonisation methods are equal in their ability to preserve biologically relevant signals, which should be considered in study design.

Taken together, our findings underscore the difficulty of selecting an optimal harmonisation method, as each approach offers some advantages and limitations depending on the analytic goal. Feature-level methods were highly effective at removing site-related variance, and were straightforward to implement, as they require only a brief preprocessing step before statistical analysis. Image-based harmonisation produced more varied results; only IGUANe reduced site-identifiability. However, they generally preserved the prediction of biologically or clinically meaningful outcomes, and in some cases even improved them, particularly for symptom predictions. In contrast to feature-level methods, image-level approaches are more complex to deploy, typically requiring computationally intensive image processing pipelines and access to raw imaging data. Nonetheless, this added effort might be worthwhile, as image-based methods offer greater flexibility for downstream analyses, as they can be used to generate harmonised images suitable for voxel-level investigations. As such, researchers must weigh both performance metrics and practical constraints when choosing the most appropriate harmonisation strategy for their specific research objectives.

Several limitations should be considered when interpreting our findings. First, harmonisation was evaluated exclusively using FreeSurfer-derived features, and results may not generalise to voxel-wise or surface-based analyses, which could be differently affected by site variance. Relatedly, it remains challenging to determine which components of biologically meaningful variance may be inadvertently attenuated by the harmonisation process. While feature-level methods appear to be highly effective at eliminating site identifiability, they may also suppress subtle but valid sources of inter-individual variability. Regarding image-level methods, HACA3 showed promise in some analyses, but was applied here only to T1-weighted images, despite being designed to leverage multi-modal inputs such as T2-weighted scans. This may potentially have limited its effectiveness. A further concern is the potential for data leakage in feature-level harmonisation pipelines (Marzi et al., 2024), which could have artificially inflated performance metrics. Additionally, our study did not include supervised harmonisation approaches, in which the same individuals are scanned across multiple sites (Zuo et al., 2023), offering a gold-standard for assessing harmonisation efficacy. Finally, the relatively modest classification accuracies observed in our analyses likely reflect the fact that we relied solely on structural imaging data, without incorporating demographic or clinical variables, and using linear prediction models without hyperparameter optimisation that might otherwise enhance predictive performance.

Future work should explore new harmonisation methods and the extension of harmonisation techniques to other data modalities (e.g., diffusion, functional MRI, magnetic resonance spectroscopy) and assess their impact at the voxel level. Finally, evaluating the effects of harmonisation on other types of downstream multivariate analyses, such as clustering or normative modelling, will also be critical to enable higher-quality multi-site neuroimaging studies of the future.

## Data Availability

The data used in the study is available via the Psychsis MRI ShareD Data Resource (psyshared.com)

https://psyshared.com/Home.html

## Psy-ShareD Partnership

Sara Bucci (King’s College London, UK), Ary Gadelha (Federal University of São Paulo, Brazil), Philip McGuire (University of Oxford and King’s College London, UK), Stefan Borwardt (University of Lübeck, Germany), Akira Sawa (Johns Hopkins University, USA), Camilo de la Fuente Sandoval (Instituto Nacional de Neurología y Neurocirugía), Paola Dazzan (King’s College London, UK), Andrew Lawrence (King’s College London, UK), Gemma Modinos (King’s College London, UK), Sukhi Shergil (King’s College London, UK), Mihai Avram (University of Lübeck, Germany), Jun Miyata (Kyoto University, Japan), Sadao Otsuka (Kyoto University, Japan), Neelabja Roy (National Institute of Mental Health & Neurosciences (NIMHANS), India), Daniel Kesser (Ludwig Maximilian University of Munich, Germany), Giacoma Cecere (University of Zurich, Switzerland), Kyle Jensen, (Georgia State University, USA), Lena Lim (Nanyang Tech University, Singapore), Albert Yang (Taipei Veterans Hospital, Taiwan), Alejandro Roig Herrero (University of Valladolid, Spain), Rafael Romero García (University of Sevilla, Spain), Benedicto Crespo (University of Sevilla, Spain), Alice Egerton (King’s College London, UK), Paolo Fusar-Poli (King’s College London, UK), Cathy Davis(King’s College London, UK), Mikkel Erlang Sørensen (CNSR, Denmark), Peter Uhlhaas (Charité - Universitätsmedizin, Germany), Russell A. Poldrack (University of California, Los Angeles, USA), Vince D. Calhoun (Georgia State University, USA).

